# Seroprevalence of SARS-CoV-2 IgG Specific Antibodies among Healthcare Workers in the Northern Metropolitan Area of Barcelona, Spain, after the first pandemic wave

**DOI:** 10.1101/2020.06.24.20135673

**Authors:** J Barallat, G Fernández-Rivas, B Quirant-Sánchez, V González, M Doladé, E Martinez-Caceres, M Piña, J Matllo, O Estrada, I Blanco

## Abstract

**Background:** The rapid spread of Severe Acute Respiratory Syndrome Coronavirus 2 (SARS-CoV-2) around the world has caused a global pandemic, infecting millions of individuals worldwide, with an unprecedented impact in health care systems worldwide. Healthcare workers are one of the risk groups that need to be well characterized due to their strategic role in the management of patients, presently and in prevention of healthcare needs for future outbreaks. This study presents the results of the first SARS-CoV-2 seroprevalence study in the Northern Metropolitan Area of Barcelona, Spain.

**Methods:** IgG SARS-CoV2 antibodies were analyzed in serum samples from 7563 healthcare workers of the Northern Metropolitan Area of Barcelona taken during the pandemia (from May 4th to May 22^nd^, 2020) by chemiluminescence assays.

**Results:** A total of 779 of 7563 (10.3%) healthcare workers had detectable anti-SARS-CoV-2 IgG (specific for either S1/S2 or N antigens). No significant differences were observed between those working at primary care or at the reference hospital.

Interestingly, in 29 (8.53%) of the previously confirmed positive reverse-transcriptase polymerase chain reaction (rRT-PCR) patients SARS-CoV-2 IgG (S1/S2 or recombinant N antigen) were negative.

**Conclusion:** Seroprevalence of anti-SARS-CoV-2 IgG in the healthcare workers of the Nord Metropolitan Area of Barcelona was significantly increased in comparison with the general population in the same geographical area. These results give us an important insight for a better understanding of SARS-CoV-2 epidemiology, in a collective that is essential for the response against this pandemic.

## INTRODUCTION

On January 2020, the World Health Organization (WHO) received information from the National Health Commission about an outbreak, which was highly suggestive to be associated with exposures in one market in Wuhan, China. Chinese authorities identified a new type of coronavirus (novel coronavirus, nCoV), which was isolated on January 7^th^, 2020. On March 11^th^, 2020, WHO declared SARS-CoV-2 a pandemic and, as of June 1^st^, 2020, more than 6 million total cases had been reported worldwide [1]. SARSCoV-2 viral RNA has been detected in both throat and nasal swabs of infected individuals by rRT-PCR. This technique is currently the gold standard for diagnosis of active infection. In most cases, viral RNA becomes almost undetectable 14 days post-illness from symptom onset, although RNA persistence for longer periods does occur sometimes [2] Additionally, IgG antibodies recognizing specific proteins of the virus do peak at 3-4 weeks after symptom onset and most individuals are able to maintain a sustained humoral response against SARSCoV-2 for at least 3 months [3].

SARS-CoV-2 constituent proteins include spike (S), envelope (E), membrane (M), nucleocapsid (N), and other ones with still unknown functions. Most exposed among those are the S proteins, being the N protein also abundantly expressed during infection [4] Both, S and N proteins are immunogenic and antibodies against them have been postulated as potential biomarkers for SARS-CoV-2 current or past infection. The S protein may also specifically mediate membrane fusion and induce neutralizing antibodies in the host, raising the possibility that certain antibodies against them may play a key role in the host response against SARS-CoV infection [5]. S comprises two functional subunits responsible for binding to the host cell receptor (S1 subunit) and fusion of the viral and cellular membranes (S2 subunit). Using both S1 and S2 reduces the possibility of cross-reaction with other coronaviruses [6].The biological function of N protein is thought to participate in the replication and transcription of viral RNA and to interfere with cell cycle processes of host cells. In addition, N protein is highly immunogenic in many coronaviruses, and abundantly expressed during infection [7], suggesting that antibodies that specifically recognize this protein might be useful in early diagnosis of the infection. Furthermore, it has been suggested an intriguing role for N protein in the primary humoral immune response against SARS-CoV infection [5].

Sero-epidemiology is a powerful tool to understand how diseases spread in specific environments and may help to design and to monitor vaccination programs [8]. By using seroprevalence surveys, we can learn about the total number of people that have been infected, including those that might have missed diagnosis. These surveys can also help to estimate the percentage of the population that has not yet been infected, helping public health officials plan for future healthcare needs.

When considering undertaking a seroepidemiological study, it is important to choose the priority public health questions to which serology can contribute: a) the antigens/antibodies to be studied; b) the populations of interest; c) the best sampling method to provide a representative sample of those populations; d) the most appropriate laboratory assays and e) how data will be managed, analyzed and reported [8].

Spain is the country with the highest number of coronavirus infections among healthcare workers, according to available official data [9]. Knowing the prevalence of infection among healthcare workers is particularly important, since their role in the pandemic implies a risk of high exposition against this pathogen. Protective measures and safety protocols have been used in order to minimize the risk of healthcare workers.Seroprevalence studies will be useful to assess the effectiveness of these protocols and to design new strategies against potential new outbreaks. A first serological study led by ISGlobal and the Hospital Clinic of Barcelona, revealed that 11.2% of the hospital staff was infected by SARS-CoV-2 [10]. The estimated SARS-CoV-2 prevalence in the general population in the area of Barcelona has been estimated to be around 7.1% [11].

The Catalan Institute of Health (ICS) is the largest public health service company in Spain and is affiliated to the Department of Health of the Catalan Government. The ICS is formed by eight hospitals and about 300 primary care teams. It provides healthcare to almost six million users, a figure that represents 75% of the total number of people with healthcare rights in Catalonia.

In the Northern Metropolitan Area of Barcelona, the ICS provides primary care services to nearly 1,400,000 citizens in 71 municipalities. In this Metropolitan Area, Germans Trias i Pujol University Hospital, in Badalona is the reference center for the high-complexity care of the 800,000 citizens of Barcelonès Nord and the Maresme and the basic general hospital of more than 200,000 residents from Badalona city and other surrounding municipalities. Primary care is provided by 66 teams and 37 support care units, working in a total of 84 Primary Care centers and 22 local clinics.

As this area of Barcelona had a high incidence of Coronavirus disease 2019 (COVID-19) cases, it was important to determine the impact of the pandemics on the health care system clusters. Hence, in this study we analyzed the SARS-CoV-2 IgG seroprevalence in Healthcare workers of the Northern Metropolitan Area of Barcelona, Spain.

## METHODS

### Study design

Healthcare workers of the Northern Metropolitan Area of Barcelona, Spain were recruited in a prospective cross-sectional study.

From May 4th to May 22^nd^, 2020, all Healthcare workers of the ICS-Northern Metropolitan Area of Barcelona (n=9315) were offered to have serum testing performed for SARS-CoV-2 IgG antibodies. This program was not offered as a research protocol but as a service to healthcare workers. According to official data [1], SARS-CoV-2 diagnosis peak in Spain was reached on April 1^st^ 2020.

The participation in this study was voluntary; healthcare workers were neither selected for participation based on symptoms nor previous exposure to COVID-19. All individuals willing to participate fulfilled a brief epidemiological questionnaire and gave permission to access their clinical records. The questionnaire included demographic data, professional information, a direct questions about if they have been diagnosed of COVID-19, or if they presented any of the most characteristic COVID-19 symptoms, such as cough, respiratory distress, fever, chills, headache, sore throat, anosmia, ageusia or asthenia.

### Laboratory analysis

Serum testing was conducted by the Regional Clinical laboratory using the quantitative SARS-CoV-2 S1/S2 IgG LIAISON® test (DiaSorin, Vercelli, Italy) on the LIAISON XL platform, following the manufacturer’s instructions. This test discriminates among negative (<12AU/mL; with 3.8 as the limit of IgG detection), equivocal (12.0 – 15.0 AU/mL) and positive (> 15.0 AU/mL) subjects. But due to recommendations provided by the same manufacturer equivocal zone was broadened to (9.0 – 15.0 AU/mL) and data were reanalyzed [12].

In those cases in which a) IgG anti S1/S2 quantification was higher than the limit of detection (i.e. >3.8 AU/mL) but did not reach the limit of discrimination (i.e. <15 AU/mL) and/or b) when the healthcare workers answered the questionnaire saying that he or she had been diagnosed of COVID-19 but IgG anti S1/S2 where lower than 15 AU/ml, aditional serological study was performed using a different antigen (N) as a target. In this case, a SARS-CoV-2 IgG test (Abbott Diagnostics, Sligo, Ireland) was run on an Architect i2000 platform. (Figure 1). This test discriminates among negative (<1.4 Index (S/C)) and positive (≥1.4 Index (S/C) subjects.

**Figure 1.**
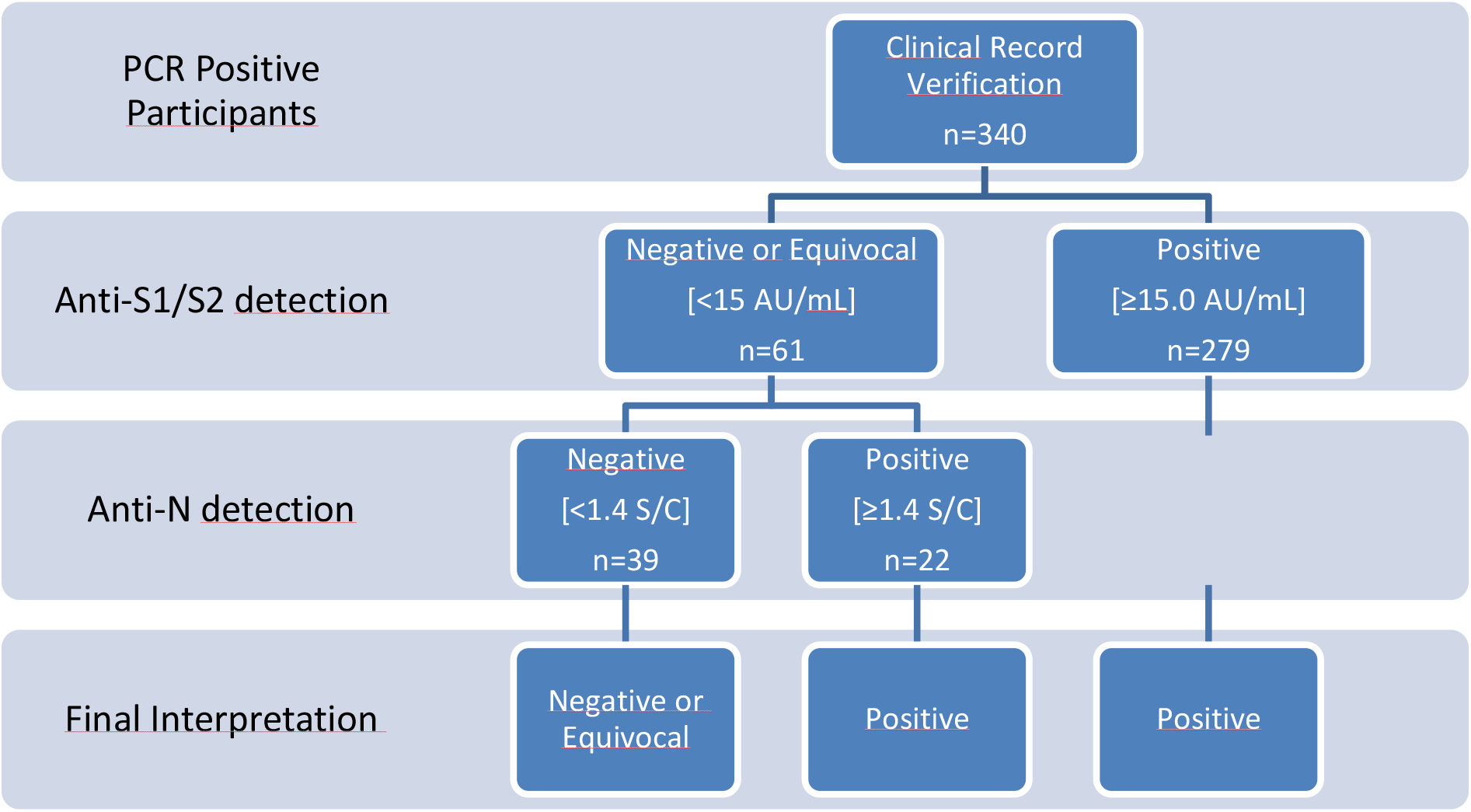
Algorithm suited in rRT-PCR participants.

Positive and equivocal results were accompanied by a statement that the results did not indicate immunity to COVID-19 and healthcare workers should continue to wear full personal protective equipment. Participants with equivocal results were offered to be retested 4 weeks after the initial sample extraction.

### Statistical analysis

Categorical variables were expressed as frequencies, while quantitative variables were expressed as the mean and standard deviation (SD). Qualitative variables were compared with Fisher’s exact test, while quantitative variables were compared using t-student test. These analyses were conducted with version 20 of SPSS.

#### Ethical considerations

As specified above, this program was not offered as a research protocol but as a service to ICS-Northern Metropolitan Area of Barcelona employees. All participants voluntarily accepted to participate and gave informed consent to review their health records. Positive and equivocal results were accompanied by a statement that the results did not indicate immunity to COVID-19 and healthcare workers had to continue wearing full personal protective equipment.

## RESULTS

A total of 7563 healthcare workers of Northern Metropolitan Area of Barcelona participated in the study. The participation rate was 81.2%.

Subjects’ characteristics are showed in Table 1. The mean age was 43.81 ± 12.43 years. 5746 participants were female (75.97 %) and 1.817 males (24.02%). From all of them, 4153 participants worked at Primary Care (54.91 %) and 3410 (45.09%) at Germans Trias i Pujol Hospital (45.09%), Tertiary Care.

**Table 1.** Demographic characteristics and antibody reactivity of participants.

| **Characteristics** | **Participants**<br>**n (%)** | **Positive SARS-CoV-**<br>**2 IgG S1/S2**<br>**n (%)** | **Positive SARS-CoV-2 IgG**<br>**S1/S2 or SARS-CoV-2 IgG N**<br>**n (%)\*** |
| --- | --- | --- | --- |
| **Entire sample** | 7563 (100) | 712 (9.41) | 779 (10.30) |
| **Sex** |  |  |  |
| **Male** | 1817 (24.02) | 182 (10.02) | 193 (10.62) |
| **Female** | 5746 (75.97) | 530 (9.22) | 586 (10.20) |
| **Age** |  |  |  |
| **18-34** | 2025 (26.8) | 209 (10.32) | 227 (11.21) |
| **35-54** | 3665 (48.4) | 315 (8.59) | 354 (9.66) |
| **≥55** | 1873 (24.8) | 188 (10.04) | 198 (10.57) |
| **Location** |  |  |  |
| **Primary Care** | 4153 (54.91) | 381 (9.17) | 415 (9.99) |
| **Tertiary Care** | 3410 (45.09) | 331 (9.71) | 364 (10.67) |
| **Health Care Job** |  |  |  |
| **Nurse** | 2246 (29.70) | 216 (9.62) | 239 (10.64) |
| **Physician** | 1838 (24.30) | 193 (10.50) | 215 (11.70) |
| **Nursing**<br>**Assistant** | 1103 (14.53) | 106 (9.61) | 116 (10.52) |
| **Health Care**<br>**Support**<br>**Services** | 349 (4.46) | 35 (10.03) | 35 (10.03) |
| **Administrative**<br>**Healthcare** | 1195 (15.80) | 77 (6.44) | 79 (6.61) |
| **Other** | 832 (11.21) | 85 (10.22) | 95 (11.42) |
| **COVID**<br>**Symptoms** |  |  |  |
| **Yes** | 3475 (45.95) | 523 (15.05) | 567 (16.32) |
| **No** | 3646 (48.21) | 129 (3.53) | 143 (3.92) |
| **N/A** | 442 (5.84) | 60 (13.57) | 69 (5.61) |
| **Previous COVID**<br>**Diagnosis** |  |  |  |
| **Yes** | 385 (5.09) | 291 (75.58) | 313 (81.30) |
| **No** | 6740 (89.12) | 362 (5.37) | 398 (5.90) |
| **N/A** | 438 (5.79) | 59 (13.47) | 68 (15.52) |
| **SARS-CoV-2**<br>**Confirmed rRT-**<br>**PCR Positive** |  |  |  |
| **Yes** | 340 (4.51) | 279 (82.06) | 301 (88.53) |
| **Hospital admitted**<br>**for COVID** |  |  |  |
| **Yes** | 59 | 46 (77.97) | 46 (77.97) |
| **No** | 7044 | 605 (8.59) | 663 (9.41) |
| **N/A** | 460 | 61 (13.26) | 70 (15.20) |

\*N antigen determination was performed in 646 participants, according to the laboratory analysis algorithm.
N/A (Not applicable).

A total of 712 out of the 7563 participants (9.41%) were positive for S1/S2 IgG; 6260 were lower than 3.80 AU/mL (82.7%). The percentage of positivity was lower in the age group of 35-54 (8.59 %). There were no differences related to sex. Administrative Healthcare workers showed the lowest percentage of positive samples (6.44%). (Table 1). In 3475 (45.95%) healthcare workers who claimed suffer COVID-19 related symptoms, 523 (15.05%) were positive for S1/S2 (Table 1).

A total of 385 (5.09%) participants answered to the questionnaire that they had been diagnosed of COVID-19, 59 of them had been hospitalized. In 340 participants, a positive rRT-PCR was confirmed reviewing the medical records. Among those, 279 (82.06%) were positive for S1/S2 IgG.

In samples from healthcare workers claiming to have been previously diagnosed of COVID-19 but S1/S2 IgG where lower than 15 AU/ml (n: 94) an additional serological test to determine anti-N was performed. Twenty two of these 94 samples (23.4%) showed positivity to IgG against the nucleocapsid antigen. Also, samples from healthcare workers in which S1/S2 IgG quantification was higher than the limit of detection (>3.8 AU/mL) but did not reach the limit of discrimination (<15 AU/mL) (n: 591) were also tested to determine N antibodies. Sixty three of these samples (10.65%) showed positivity to N IgG.

Taking into account the detection of anti-N IgG antibodies, the seroprevalence of the entire sample increased to 10.3% (n: 779/7563). Accordantly, the percentage of positive SARS-CoV-2 IgG in Healthcare Workers with a previous positive rRT-PCR increased after analyzing the presence of IgG anti-Nucleocapsid, finally been of 88.53% (301/340). However, Thirty-five of these Healthcare Workers having a previous positive rRT-PCR did not show discriminable levels of antibodies with any of the tests.

We recalculate all data according with the suggestions of Bonelli et al [12] increasing the range of equivocal results for LIAISON SARS_CoV S1/S2 from 9 – 15 AU/ml. As expected, the percentage of positive samples did not change but the number of negative samples of the entire series decreases to 88.39 % (6685/7563). Furthermore, only 29 (8.53 %) of the 340 rRT-PCR-diagnosed patients had non-discriminable levels of IgG either anti-S1/S2 or N antigens.

## DISCUSSION

Serological assays play an important role in the knowledge of the impact and evolution of the COVID-19 pandemic, especially in healthcare workers. In our opinion these data are useful to answer two questions: 1) the exposure of our healthcare workers to SARS-CoV-2 during the crisis and 2) the analysis of the humoral response in a well characterized population.

To know the seroprevalence of our healthcare workers compared to our local population is important to assess the efficiency of the safety protocols established to protect our colleagues. Spain is known to have established a strict lockdown in the country’s general population since the pandemic was declared on March 13, 2020. Consistently, a higher seroprevalence among Healthcare workers was expected, since this collective has been in contact with infected patients during more than three months, in an outstanding professional task. SARS-CoV-2 IgG seroprevalence in the Barcelona area has been estimated to be at around 7.1%, data obtained, interestingly, in an almost perfect timing match with our study [10]. Our results among healthcare workers were slightly higher, but not as concerning as they could have been expected without the application of strict protocols.

In relation to the analysis of the humoral response, the study of a whole population of individuals comprising all types of cases, including mild forms, can be very useful, since most published serology data are referred to hospitalized patients. Serological data of rRT-PCR positive but mild symptomatic patients are scarce [13] and this information is important for a better understanding of this infection. Our data showed that 8.53% of diagnosed patients tested negative for IgGs against SARS-CoV-2. Thirty-nine of the Healthcare workers with positive rRT-PCR required hospitalization. Only, one of the hospitalized did not showed discriminative SARS-CoV-2 IgG either S1/S2 or anti-nucleocapsid. This patient was clinically classified as a mild COVID-19. These results underline the relevance of characterizing broad cohorts of patients, and not only the ones with most relevant clinical manifestations.

Few seroprevalence studies of healthcare workers have been published so far and those have various outcomes depending on diverse factors, others than those specific for the analytical techniques. The seroprevalence of 316 healthcare workers of a tertiary hospital in Germany was 1.6% [14]. Another study in Italy, a country with also a high burden of COVID-19 performed with the same platform showed similar data to the ones of the present study (447 positive/3985 participants; 11%) [15]. In Barcelona, another study led by IS Global from 583 workers at the Clinic Hospital gave a seroprevalence of 11.3% considering a pool of antigens and the three immunoglobulin isotypes (IgG+ IgA+ IgM) using an in house test based on Luminex platform [10]. Our results analyzing only IgG specific levels has very similar results to the last one.

A total of 3646 participants answering the epidemiological survey claimed to be fully asymptomatic. Among those, 143 (3.92%) did test positive for IgG anti SARS-CoV-2. As social restrictions are being eased, characterizing asymptomatic infected individuals is crucial to understand how the disease is spreading [16]. As our cohort of subjects is notably large, we cannot discard that a small proportion of these results could be false positives. In this context, our primary IgG test, anti S1/S2, claims a specificity of 98.5%. Even taking these values into account, there is still a high likelihood that most of the asymptomatic individuals that tested positive were infected.

In our study, we observed that 8, 53% of the individuals that theoretically had passed COVID19 infection did not present a discriminative level of IgG using two different groups of antigens (S and N). Any of both methods do not guarantee a 100% analytical sensitivity. LIAISON® SARS-CoV-2 S1/S2 IgG displays a sensitivity of 86.8-99.5%, according to the manufacturer. Abbott Architect IgG claims a sensitivity of 95.89-100%, but external evaluations have reported it to be around 93% and dropping along the days of evolution of the illness [17]. Besides, other explanations can arise. The first one could be that some individuals that reported symptoms or previous rRT-PCR positive were not really infected. This explanation was discarded after reviewing their personal records. A second possibility could be that samples were obtained during the first 10 days of infection, when it is well known that most of the IgG are not detectable [3]. This was also discarded as all those patients had positive rRT-PCR from at least 23 days before the serum sampling. Another possible explanation would be that infection occurred, but the innate immune system eliminated the virus, not allowing to organize a relevant specific response.Finally, there are a percentage of individuals (1-2%) [14] who do not show detectable antibodies for unknown reasons.. All this serves as a reminder that individual protective measures should never be discontinued, regardless of symptomatology.

Another point of discussion is the putative protective value of antibodies against reinfection. Even considering tests as highly sensitive, much about protective immunity is unknown [6][7], however it seems that spike glycoprotein antibody tests will be preferred as a target for further studies related with the neutralizing antibodies [13].

Our study has several limitations. No other immunoglobulin isotypes were analyzed, and it cannot be discarded that some individuals might be positive for IgA or IgM and the moment of blood extraction. Moreover, not all antigens have been tested in all individuals. Only those with a report of disease or with detectable but non-positive SARS-CoV-2 IgG S1/2 were analyzed for both N and S antigens.

Nevertheless, some strong points can be considered in the current study. First, the large number of individuals tested in a clearly restricted sanitary area suffering a high impact of the pandemic. Second, the study set-up, in the context of a high throughput diagnostic laboratory, showing the technical viability of testing high number of patients in a short period. Third, the concordance of the results with reported infection and/or rRT-PCR results from the staff.

Interestingly, this study opens new research perspectives, as it has identified a group of individuals in which, despite of having suffered COVID19 infection, not detectable antibodies were found. Further analysis considering broader antibodies isotypes as well as cellular responses need to be implemented in routine bases to better characterize these population.

In summary, we report that seroprevalence of anti-SARS-CoV-2 IgG antibodies in the healthcare workers of the Nord Metropolitan Area of Barcelona gives was slightly increased in comparison with the general population in the same geographical area and similar to other referent hospitals in Barcelona. Interestingly a similar prevalence was observed in Primary care and Hospital workers and no differences were observed in between de healthcare work positions.

## Data Availability

All data referred to in the manuscript or any questions related with the manuscript can be consulted throughout the corresponding author.

## Acknowledgments

The authors would like to thank all participants and to all nurses and laboratory technicians involved in this study, for their help in specimen collection, specimen processing and for their outstanding work during this pandemic.

